# Knowledge of self-care and complications of diabetes mellitus among diabetes mellitus patients in Addis Ababa, Ethiopia

**DOI:** 10.1101/2023.11.17.23298674

**Authors:** Roman Getachew, Dagmawit Tewahido

**Affiliations:** Epidemiology and Biostatistics Department, Addis Continental Institute of Public Health, Addis Ababa, Ethiopia; Nutrition and Behavioral Sciences Department, Addis Continental Institute of Public Health; Addis Ababa, Ethiopia

## Abstract

**Background:** Diabetes mellitus is a progressive disease that compromises the quality of life of the victims. Patients’ knowledge about diabetes mellitus self-care is vital in achieving targeted glycemic control and minimizing complications of the disease. However, there are limited studies in Ethiopia on the subject to guide interventions. Hence, this paper aimed to assess knowledge about diabetic self-care and complications among diabetes mellitus patients in Addis Ababa, Ethiopia.

**Methods:** An institutional-based cross-sectional study design was employed. A structured questionnaire was used to assess knowledge; participants who responded correctly to at least 6 of the eight questions were classified as having good knowledge about the complications and those who answered <6 questions correctly were classified as having poor knowledge. Participants who correctly responded to ≥30 of the 37 self-care questions were classified as having good knowledge about diabetes self-care; those who correctly responded to <30 questions were classified as having poor knowledge. Multivariate logistic regression analyses were used to identify associated factors.

**Result:** Overall, 202 (51.7%) of the study participants had good knowledge about the complications, and 279 (71.4%) of the participants had a good knowledge of self-care. Receiving diabetic education was a significant modifiable factor for having poor knowledge about both the complication (AOR= 3.078(1.323,7.160)) and the self-care (AOR=7.645(3.153,18.538)).

**Conclusion:** About half (51.7%) of the participants had good knowledge about the complications, and about three in four (71.4%) had good knowledge about self-care. Receiving diabetic education was significantly associated with both complication and self-care knowledge status. Focusing on the quality and quantity of the information that is delivered to diabetes mellitus patients can improve the quality of care and the outcome.

## Introduction

Diabetes is one of the non-communicable chronic metabolic disorders characterized by hyperglycemia progressively leading to complications to organ systems (1,2). Such complications have been becoming more prevalent globally particularly in developing countries (2). From a study in Ethiopia, the overall prevalence of Diabetes Mellitus (DM) complications is 72.4% in Type 2 diabetes mellitus patients and 27.6% in Type 1 diabetes mellitus patients (3) Knowledge about the complications and self-care has significant effect on adequate glycemic control and treatment success(4–6) and is one of the patient related factors (7).

Different studies, however, show that complications of diabetes are not well known by the patients themselves. In a study from Nepal, good knowledge about the complications was only 24.2% (5). In China and Saudi Arabia, only 61.8% and 45.5% of participants knew about retinopathy (8,9). Knowledge about the complications was 65% in Senegal (10), 45.9% in Northern Ghana (11) and only 13.1% in Sampa, Ghana (12). In studies from North West Ethiopia and Addis Ababa, 48.5% and 54.9% of good knowledge was recorded respectively (13,14).

Similarly limited knowledge about diabetic self-care was recorded to be 58.28% and 76.6% in Jordan and Saudi Arabia (15,16), 79.5% in Nigeria (17), 67.8%, 61.3%, 70.4% and 63.3% in studies conducted in North Shewa, Dessie hospital, Ayder hospital and Jimma medical center, Ethiopia, respectively (18–21).

Different factors were observed to positively or negatively affect the knowledge that DM patients acquire. While good self-care is associated with being employed, urban residency, receiving information on diabetic self-care, being married, male gender and earning medium income, good knowledge about the complications of the disease is associated with-with male gender, younger age, higher financial status, higher literacy level, and longer duration of illness (12,14,22,23).

Knowledge that DM patients have about the disease, it’s complications and the self-care is vital in achieving good glycemic control. Patient education and awareness creation programs are core in equipping DM patients and those who are involved in giving care with the right knowledge. But there are concerns about the success of these activities as it can be seen through the burden of the increasing prevalence of the complications and the failure to achieve the goals set by nations and the world. There are also limited studies on the subject hence this study was conducted with the aim to assess knowledge about complications and self-care and the associated factors.

## Methodology

### Study design

Institutional based cross-sectional study design was used to assess the knowledge of diabetes mellitus patients regarding complications and self-care of the disease.

### Study setting

The study was conducted at Minilik II referral hospital which is among the 7 public hospitals under Addis Ababa city administration Health Bureau. The hospital gives service for patients with communicable and non-communicable diseases such as diabetes mellitus. DM related services include screening and diagnosis, treatment and health education. The diagnosis and treatment including admissions are given as per the standard cost of the hospital where as health educations are given at a mass for free in addition to the individualized health education given at different points including at inpatient wards. Clients who are embraced under community-based health insurance also get the service.

### Study population

All types of adult DM patients who came to medical referral clinic between March 17 and April 20 were potential candidates. Those who were more than 18 years old, those who were on treatment for at least one year, those who were able to respond verbally and those who were not diagnosed to have mental illness were included given their willingness and those who were identified to have critical physical and mental illness during interview and those who were admitted at time of interview were excluded.

### Operational definitions

**Diabetic education-** Diabetic education includes provision of information mainly about diet, exercise, foot care, cessation of smoking, blood glucose monitoring and medication that DM patients should be aware of (24). This study focuses on the diabetic education given at mass level, at the outpatient clinics and the individualized education that patients receive from their respective physician.

### Sampling and Sample size

Single and double population proportion formulas were used to get a sample size of 420 including the 10% non-responsive rate. Systematic random sampling was employed to recruit participants. The first patient was selected by lottery method and every 5^th^ of them were interviewed.

### Data Collection

A structured and pre-tested questionnaire was developed from conducting literature review and taking other studies experiences (19,21,25) for the self-care related knowledge questions and from studies in Ghana(12,22) for the complication related knowledge questions with some modifications (26,27). The questionnaire had four sections. Section one included demographic related questions; section two clinical condition of patients; section three questions related to knowledge about the complications; and section four questions on the self-care. Each question had “yes”, “no” and “I don’t know” answer options. Those with a score of >mean+1 SD were classified to have good knowledge whereas those with a score of <mean-1 SD were classified to have poor knowledge (22). Interviews were conducted face-to-face in Amharic (the national language) in a private space, before patients visit their physicians. Pre-test was conducted on 5% of DM patients. Data collectors were 2 nurses who were recruited from other unit of the hospital to avoid responder bias. Training was given for data collectors for one day prior to start of data collection. Supervision and monitoring were conducted regularly by the principal investigator.

### Data analysis

Data were collected and cleaned using Kobo Toolbox and exported to SPSS version 20. Analysis and conclusion were made for the 391 responses making the response rate of the study to be 93.09%. Binary logistic regression was done and those variables with P<0.02 were analyzed by multivariable logistic regression to select significantly associated factors. Hosmer and Lemeshow goodness-of-fit-test statistics were used to assess the fitness of the model (p-value ≥0.05). Multicollinearity was checked by using cut-off point based on variance inflation factor (VIF) < 10 or tolerance test > 0.1. Then, multivariable logistic regression analysis at P value < 0.05 and adjusted Odds Ratio (AOR) with 95% CI was used to measure the degree of association between independent variables and outcome variable.

### Ethical approval

Ethical approval was obtained from the review board of Addis Continental Institute of Public Health (ACIPH-MPH/039/14) and review board of Addis Ababa Health Bureau (A/A/H/12128/227). A permission letter was sought from the hospital (5503/69/14). Informed oral consent was obtained from each participant. Oral consent was approved by the ethics committee considering the study is minimal risk and anticipating lower literacy level in the old age. Participation in the study was based on free will and not related to any of participants’ regular care.

## Results

### Socio-demographic characteristics

Most of the participants-(53.2%) were males. The mean (SD) age was 57 (8.97) years and nearly 2/3 (65.2%) of the participants were over the age of 55years. Majority (43.5%) attended only primary education. Almost 99% (98.5%) of the participants are urban residents. More than 80% (83.6%) were married. Nearly 2/3 (62.4%) of the participants were unemployed. Almost 50% (47.1%) of the participants have known their diabetic status in the past 5 to 10years. Nearly three fourth (71.4%) had no known family history of DM.

**Table 1:** Socio-demographic characteristics of participants

| Variable |  | Frequency (N=391) | Percent (%) |
| --- | --- | --- | --- |
| Gender | Male | 208 | 53.2 |
|  | Female | 183 | 46.8 |
| Mean age (SD) | 57.23 (8.9699) |  |  |
| Age | 23- 45 | 38 | 9.7 |
|  | 46- 55 | 98 | 25.1 |
|  | 56- 85 | 255 | 65.2 |
| Place of residence | Urban | 385 | 98.5 |
|  | Rural | 6 | 1.5 |
| Educational level | Unable to read and write | 21 | 5.4 |
|  | Primary education | 170 | 43.5 |
|  | Secondary education | 106 | 27.1 |
|  | Tertiary education | 94 | 24.0 |
| Marital status | Married | 327 | 83.6 |
|  | Non- married | 64 | 16.4 |
| Working status | working | 98 | 25.1 |
|  | Non- working | 293 | 74.9 |
| Years since diagnosis of DM | < 5 years | 59 | 15.1 |
|  | 5 to 10 years | 184 | 47.1 |
|  | > 10 years | 148 | 37.9 |
| Family history of DM | Yes | 1 | 0.3 |
|  | No | 279 | 71.4 |
|  | I don't know | 111 | 28.4 |

### Clinical condition of participants

In this study 230 (58.8%) of the participants had one or more complications of DM and 56 (14.3%) had other chronic illnesses than DM. About 318 (81.3%) of the participants had glucometer at home but only 317 (81%) of them measure their blood sugar level. 353 (90.3 %) of participants received diabetic education of which in 94% of the cases, health professionals were the source of information. However, the health education was given only “sometimes” in 88% of the cases. Majority of the participants (44%) were taking only insulin at the time of this study.

**Table 2:** Characteristics of clinical condition of participants

| Variables |  | Frequency | Percentage (%) |
| --- | --- | --- | --- |
| Current medication | Insulin | 172 | 44 |
|  | Oral medications | 157 | 40.2 |
|  | both | 60 | 15.3 |
|  | Other | 2 | 0.5 |
| Complication of DM | Yes | 230 | 58.8 |
|  | No | 161 | 41.2 |
| Any other chronic illness | Yes | 56 | 14.3 |
|  | No | 335 | 85.7 |
| Presence of glucometer at home | Yes | 318 | 81.3 |
|  | No | 73 | 18.7 |
| Frequency of blood glucose measurement at home | Never | 74 | 18.9 |
|  | Sometimes | 309 | 79.0 |
|  | Often | 8 | 2.0 |
| Membership of Ethiopian diabetic association | Yes | 7 | 1.8 |
|  | No | 384 | 98.2 |
| Family support of any kind | Yes | 353 | 90.3 |
|  | No | 38 | 9.7 |
| Ever received diabetic education | Yes | 353 | 90.3 |
|  | No | 38 | 9.7 |
| Source of information(N=353) | Health professional | 332 | 94.1 |
|  | Media | 19 | 5.4 |
|  | Other | 2 | 0.5 |
| Frequency of diabetic education | Never | 38 | 9.7 |
|  | Rarely | 3 | 0.8 |
|  | Sometimes | 345 | 88.2 |
|  | Often | 5 | 1.3 |

### Knowledge about complications and self-care of DM

An overall good knowledge about complications of DM was recorded among 202 (51.7%) participants with a mean (SD) knowledge of 6.69 (1.4). Retinopathy, diabetic foot ulcer and hypertension were the top 3 well known complications (97.4%, 96.9%, and 96.2% respectively). Over 90% of participants were observed to know nephropathy, neuropathy and heart disease (94.4%, 91.8% and 94.6% respectively) whereas hypo sexual dysfunction was the least known complication (46.8%) by the participants.

Good self-care knowledge was recorded among 279 (71.4%) of participants in the present study with a mean (SD) of 5.939 (1.214). About 1/3 (63.9%) of participants had good knowledge about signs and symptoms of hyperglycemia. Though over 70% (72.9%) of the participants had good knowledge about signs and symptoms of hypoglycemia, only 1/3 (32.7%) of them knew about its treatment. Especially participants were seen to lack knowledge about going to health care institute when hypoglycemia occurred. Over half (57.8%) of the participants had good knowledge regarding foot care however only 50% (50.4%) of them knew about annual diabetic foot check-up at clinic level. Nearly 95% (94.6%) of participants knew about eye care and its use. Most of participants (98.2%) had good knowledge about exercise in DM patients. All participants knew that DM patients should take their medication properly and stick to the recommended diet.

### Factors associated with good knowledge about Complications and Self-care

Having good knowledge about the complications was seen to be affected by different factors; although Bivariate analysis showed that male gender, higher educational status, having job and receiving diabetic education are significantly associated with having good knowledge about the complications, multivariate logistic regression analysis revealed that only being married (AOR=2.671(1.381,5.164)), having a job (AOR=2.435(1.325,4.476)) and receiving diabetic education (AOR= 3.078(1.323,7.160)) are statistically associated with having more than 2 times better knowledge about the complications of the disease.

**Table 3:**
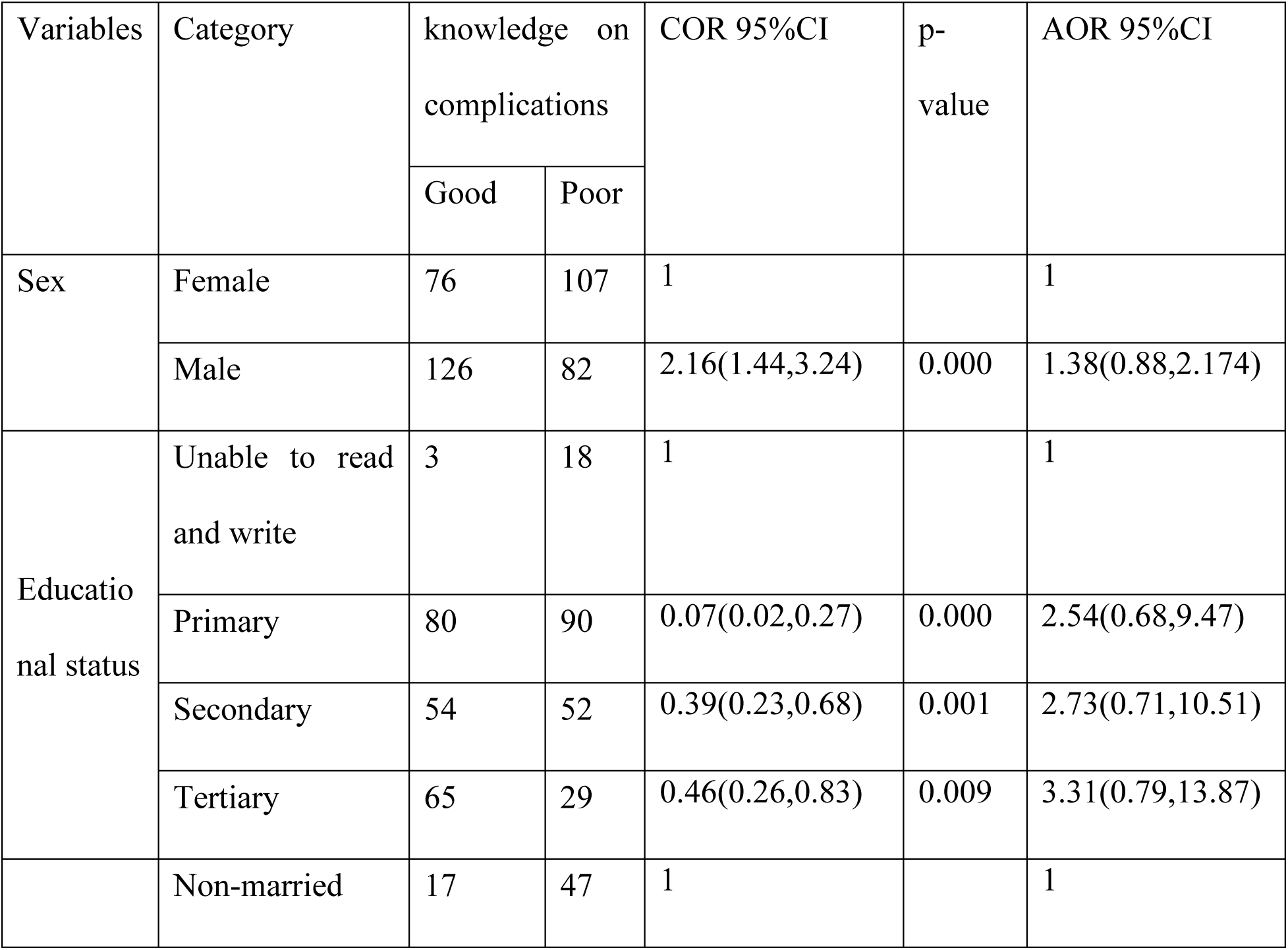
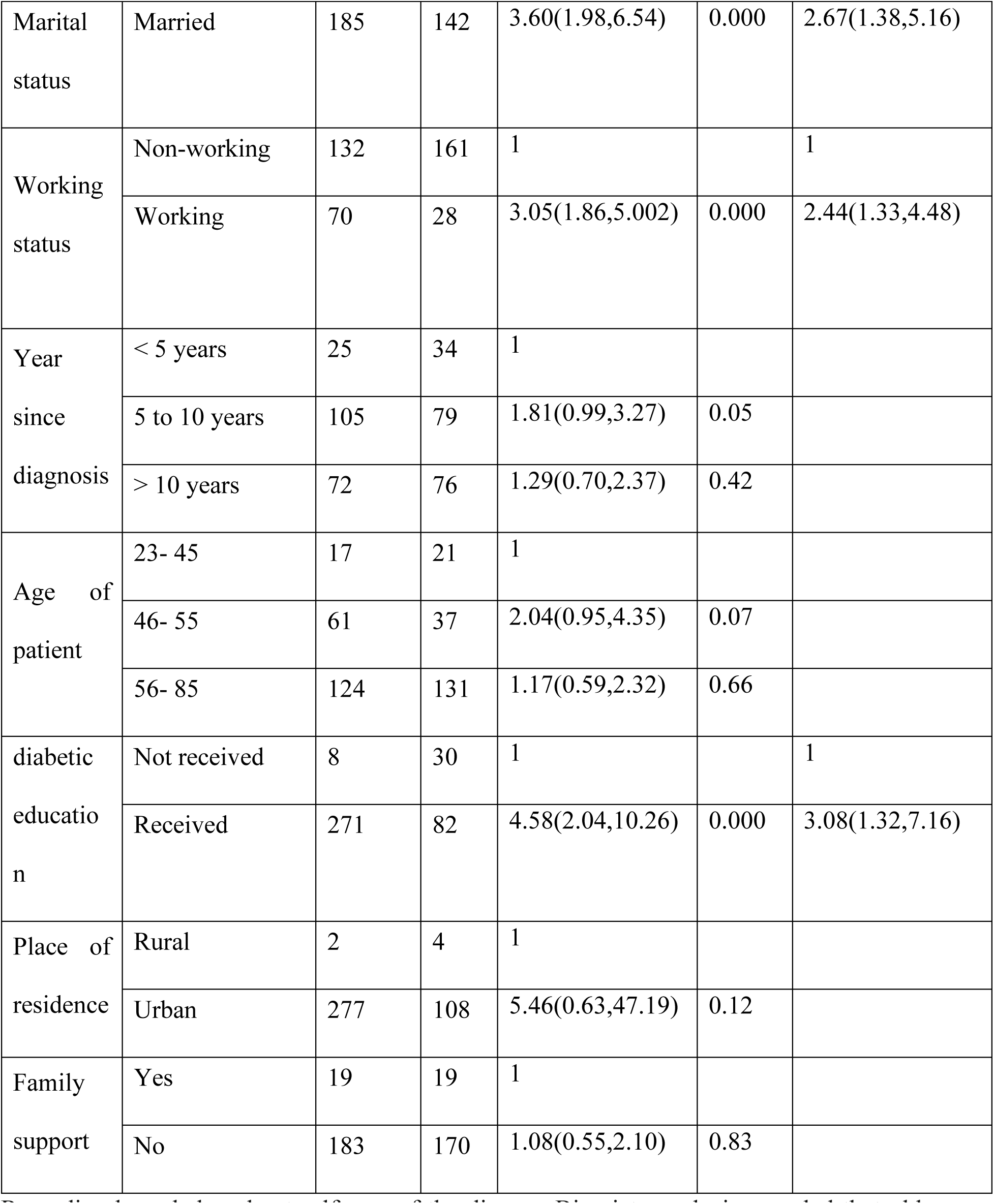
Factors associated with knowledge about complications of DM

Regarding knowledge about self-care of the disease, Bivariate analysis revealed that older age, longer duration of having the disease, receiving diabetic education and receiving family support of any kind are significantly associated with having good self-care knowledge. However, multivariable logistics regression showed that those who are 56 years of age and above have 2.76 times better knowledge than those who are 45 years of age and below (AOR=2.760(1.130,6.744)) and those who are 46 to 55 years of age have 3 times better knowledge (AOR= 3.007(1.187,7.616)) than age groups who are and below 45 years of age; those who have been with the disease for more than 10years (AOR=3.376(1.502,7.585)) and 5 to 10 years (AOR=3.833(1.776,8.272)) have more than 3 times better knowledge than those who have the disease for less than 5 years; those who received diabetic education had more than 7 times better knowledge (AOR=7.645(3.153,18.538)) about the self-care components of the disease than who didn’t receive the diabetic education.

**Table 4:** Factors associated with knowledge about diabetic self-care

| Variables | Category | Knowledge on self-care |  | COR 95%CI | p-value | AOR 95%CI |
| --- | --- | --- | --- | --- | --- | --- |
|  |  | Good | Poor |  |  |  |
| Sex | Female | 125 | 58 | 1 |  |  |
|  | Male | 154 | 54 | 1.32(0.85,2.05) | 0.21 |  |
| Educational status | Unable to read and write | 11 | 9 | 1 |  |  |
|  | Primary | 130 | 40 | 2.955(1.17,7.46) | 0.02 |  |
|  | Secondary | 72 | 34 | 1.925(0.75,4.97) | 0.17 |  |
|  | Tertiary | 66 | 28 | 2.143(0.82,5.62) | 0.12 |  |
| Marital status | Non-married | 40 | 24 | 1 |  |  |
|  | Married | 239 | 88 | 1.63(0.93,2.86) | 0.09 |  |
| Occupation | Non-working | 206 | 87 | 1 |  |  |
|  | Working | 73 | 25 | 1.23(0.73,2.07) | 0.43 |  |
| Year since diagnosis | < 5 | 19 | 40 | 1 |  | 1 |
|  | 5 to 10 | 146 | 38 | 8.09(4.21,15.53) | 0.000 | 3.83(1.78,8.27) |
|  | >10 years | 114 | 34 | 7.06(3.62,13.75) | 0.000 | 3.38(1.50,7.59) |
| Age in years | 23- 45 | 13 | 25 | 1 |  | 1 |
|  | 46- 55 | 73 | 25 | 5.62(2.49,12.62) | 0.000 | 3.01(1.19,7.62) |
|  | 56- 85 | 193 | 62 | 5.99(2.89,12.41) | 0.000 | 2.76(1.13,6.74) |
| Diabetic education | Not received | 8 | 30 | 1 |  | 1 |
|  | Received | 271 | 82 | 12.39(5.47,28.09) | 0.000 | 7.65(3.15,18.54) |
| Place of residence | Rural | 2 | 4 | 1 |  |  |
|  | Urban | 277 | 108 | 5.13.(0.93,28.42) | 0.06 |  |
| Family support | No | 16 | 22 | 1 |  | 1 |
|  | Yes | 263 | 90 | 4.02(2.02,7.99) | 0.000 | 1.34(0.55,3.26) |

## Discussion

In this study 51.4% of the participants had good knowledge about complications of DM and 73.1% about self-care. Diabetic education has been a significant factor that influenced the knowledge about both diabetic complications and the self-care. Whereas having job and being married predicted the knowledge about complications of diabetes mellitus, older age and longer duration of diagnoses were identified to positively affect knowledge about the self-care.

The overall good knowledge about the complications is higher in the present study than studies in Sampa hospital, Ghana (12), Northern Ghana (22) and Northwest Ethiopia (28) where the overall knowledge level was good among 13.1%, 45.9% and 48.5% of the participants respectively. This result can be attributed to the higher number of participants with better educational status and higher urban residential status noticed in the present study. Similar knowledge level of participants was recorded in studies conducted in Nepal (51%) (29). But the present study has revealed a lower overall good knowledge level than conducted in Addis Ababa (54.9%) which can be due to the presence of lower number of participants who attended primary and secondary school when compared to those who didn’t attend at all in the present study (14).

Having job, being married and receiving diabetic education are related with good self-care knowledge. Despite the type, participants who have a job are found to acquire 2 times better knowledge than those who are not working (AOR=2.44(1.33,4.48)) and those who are married have nearly 3 times better knowledge than those who are not married (AOR=2.67(1.38,5.16)). Those patients who received diabetic education or participated in diabetic counseling sessions had 3 times more knowledge (AOR=3.08(1.32,7.16)) than those who didn’t receive diabetic education which is supported by the studies from Nepal (29) and Addis Ababa (14). Age, educational level, duration of DM, and family history of DM are found to be predictors of knowledge level about the complications of DM in other studies unlike the present study.

Regarding knowledge of participants about self-care, the overall good knowledge recorded in this study was higher than those conducted at Jimma medical center, Ethiopia (63.3%) (21), North Shewa Ethiopia (67.8%) (18) and South-west Ethiopia (64.8%) (30). These results could be due to the lower number of participants who are unable to read and write and higher urban living status in the current study. However, a similar knowledge level was recorded in a study at Ayder hospital Ethiopia (70.4%) (25).

In this study older age, longer duration of diagnosis and receiving diabetic education has been found to be protective for having good knowledge about the self-care of the disease. Thos who are in the age group of 46 to 55 years of age showed a 3 times better knowledge (AOR=3.007(1.19,7.62)) and those who are 56 to 85 years of age recorded a 2.8 times better knowledge (AOR=2.760(1.13,6.74)) than those who are in the age group 23 to 45 years of age. Participants who have been diagnosed in the last 5 to 10 years were found to acquire 3.8 times better knowledge (AOR=3.833(1.78,8.27))and those who had the disease for over 10years had 3.4 times better knowledge (AOR=3.376(1.50,7.59)) than those who have the disease for less than 5 years. Diabetic education also resulted in 7.6 times better knowledge in those who received the education or awareness (AOR=7.645(3.15,18.54)) than those who didn’t. Unlike the present study, in a study from Jimma medical center (21) age group of 40 to 60 years was associated to poor knowledge and duration of diagnosis less than 5 years was associated with good knowledge about the self-care. Receiving diabetic education has also been a significantly associated factor for having good self-care knowledge in a study from North Shewa Ethiopia (18)which is in line with this study. A higher knowledge about the self-care was recorded in Nigeria (79.5%) with a possible explanation to be higher number of participants who attended tertiary education (17)

Knowledge which diabetes mellitus patients have about the complications, the self-care and the disease in general has been mentioned as an important factor from different perspectives in the progress of the disease (7). It is evidenced that attending diabetic education affects knowledge positively modifying quality of life and the success in achieving good glycemic control (2,7,31–35). The present study as well as studies conducted in Ethiopia, Ghana, Brazil, Bangladesh and Nepal reviled the same effect (2,10,12,14,22,29).

Non communicable diseases are commonly considered as the problem of the developed world and the rich by the community. Intense health education and promotion as well as diabetic education are required for those who are suffering from the disease and the people who are involved in giving care. Improving the knowledge status not only of the diabetes mellitus patients and care givers but also the community at large, can create a holistic and supportive environment. It can also be the starting point in improving the burden on the health system and achieving the sustainable development goal-3 (1).

### Limitations of the study

Our study was conducted in the capital city of Ethiopia where access to education is better. Thus, the knowledge level in less urban and less educated communities in Ethiopia can be poorer than what is reported in this study therefor conducting similar studies at different settings especially at rural areas, where access to media and good health care is low, might reveal different knowledge level of diabetes mellitus patients and related affecting factors at the respective context which will contribute to act accordingly.

### Conclusion and recommendation

About half (51.7%) of the participants had good knowledge about the complications, and about three in four (71.4%) had good knowledge about self-care. Receiving diabetic education was significantly associated with both complication and self-care knowledge status. Focusing on the quality and quantity of the information that is delivered to diabetes mellitus patients can improve the quality of care and the outcome.

## Declaration

### Consent for publication

#### Availability of data and material

The datasets generated/or analyzed during the current study are available from the corresponding author on reasonable request.

#### Funding

This research has not received any funding.

#### Authors’ contributions

Dr Roman: initiated the research, conducted the research, data entry and analysis, and wrote the first draft of the manuscript. Dr Dagmawit: contributed to the supervision and advise of the research activity and write-up of the manuscript.

## Data Availability

all data produced in the present study are available upon reasonable request to the authors but are on process to link with repositories

## Acknowledgements

We thank the patients and the hospital team for their heartily cooperation.

